# A Multi-Polygenic Risk Score Approach Incorporating Physical Activity Genotypes for Predicting Type 2 Diabetes and Associated Comorbidities: A FinnGen Study

**DOI:** 10.1101/2025.09.30.25336952

**Authors:** Elina Vettenterä, Laura Joensuu, Katja Waller, FinnGen, Elina Sillanpää

## Abstract

**Aims/hypothesis:** Genetic prediction of type 2 diabetes risk has proven difficult using current methods. Recent studies have shown that genetic variants associated with physical activity behavior are linked to type 2 diabetes incidence. This study investigated how a polygenic risk score (PRS) for type 2 diabetes relates to the incidence of type 2 diabetes and its comorbidities and whether incorporating genetic risk from physical activity-related traits and measured lifestyles improves prediction. We hypothesized that adding physical activity genotypes into prediction models would improve predictive accuracy.

**Methods:** PRSs were calculated for 279,373 Finns in the FinnGen cohort (average age 62 years, 52% women). Cox proportional hazards models were used with follow-up from birth. In addition, we assessed whether predictive ability (concordance index) improved when PRSs for physical activity, sedentary time, cardiorespiratory fitness, muscle strength, and body mass index were included alongside the type 2 diabetes PRS. Finally, we assessed how smoking and body mass index changed the model’s predictive ability.

**Results:** Each standard deviation unit increase in the type 2 diabetes PRS was associated with an 8% higher risk of developing type 2 diabetes. Among individuals with type 2 diabetes, the PRS was linked to higher risks of comorbidities: 4% higher for nephropathy and retinopathy and 5% for severe cardiovascular disease, but not neuropathy. Physical activity-related PRSs were also independently associated with the risk of type 2 diabetes—lower risk for physical activity (7%), cardiorespiratory fitness (6%), and muscle strength (4%) and higher risk for sedentary time (14%) and body mass index (35%). However, physical activity-related PRSs did not significantly improve the model’s concordance index (0.644 before vs. 0.672 after adding all other PRSs). In contrast, including body mass index and smoking status increased predictive ability (c-index 0.744).

**Conclusions and applicability:** PRSs for type 2 diabetes and physical activity-related phenotypes independently predict the incidence of type 2 diabetes and comorbidities. However, adding physical activity-related scores to the model does not significantly improve prediction beyond the type 2 diabetes score. Notably, the PRS for body mass index was better than the PRS for type 2 diabetes in predicting type 2 diabetes incidence. These findings support the hypothesis that genetic pleiotropy may partially explain associations between type 2 diabetes and physical activity behavior.

**Summary boxes:** *What is already known about this subject?:*

- Genetic factors contribute substantially to the risk of type 2 diabetes, but it is primarily a multifactorial condition in which modifiable lifestyle factors—including physical activity—play a critical role in onset and progression.
- Genetic variants related to physical activity behavior have been associated with type 2 diabetes.
- The clinical utility of polygenic risk scores in predicting type 2 diabetes risk remains limited, as they explain only a small proportion of genetic variance and provide minimal improvement in risk prediction beyond established clinical risk factors. What is the key question?

- Can the risk estimates for type 2 diabetes and its comorbidities be improved by incorporating genetic risk factors associated with physical activity and measured lifestyle behaviors? What are the new findings?

- Polygenic risk scores for both type 2 diabetes and physical activity-related phenotypes independently predict the incidence of type 2 diabetes and its comorbidities.
- Incorporating physical activity-related polygenic risk scores into the model does not significantly improve predictive accuracy beyond the type 2 diabetes risk score alone.
- The findings support the hypothesis that genetic pleiotropy may partially explain associations between type 2 diabetes and physical activity behavior. How might this impact on clinical practice in the foreseeable future?

- Although polygenic risk scores for type 2 diabetes may aid in identifying high-risk individuals for targeted prevention, their integration into clinical practice requires further validation.

## INTRODUCTION

Diabetes is one of the fastest growing diseases worldwide and is projected to affect 783 million adults by 2045 [1]. This rise is largely driven by 2 diabetes, which accounts for 80–90% of all diabetes cases [2]. Type 2 diabetes and its comorbidities, including cardiovascular disease, stroke, kidney failure, and lower limb amputation, cause considerable human suffering and place a heavy burden on healthcare systems worldwide [1–3]. Globally, the main drivers of this trend are an aging population, sedentary lifestyles, and unhealthy lifestyles associated with obesity [1]. Identifying individuals at high risk of developing type 2 diabetes is thus a key component of population-level prevention strategies.

The etiology of type 2 diabetes is multifaceted, involving complex interactions between genetics, environment, and lifestyle, with effects accumulating over the life course [4]. Twin models suggest that genetics accounts for a considerable amount (30–70%) of the variation in disease risk [5,6]. Recent advancements in genetics and large genome-wide association studies (GWASs) have identified many common genetic variants that increase type 2 diabetes risk [7–9]. These findings have enabled the development of polygenic risk scores (PRSs), which estimate an individual’s genetic liability for different diseases [10]. PRSs are regarded as useful tools for predicting genetic susceptibility to complex traits [11], and for type 2 diabetes, they have been validated across populations [12,13]. They also show promise as screening tools for identifying individuals at elevated risk [13], potentially enabling more targeted preventive lifestyle interventions. In addition to identifying disease risk, genetic information may help predict the risk of type 2 diabetes comorbidities and treatment responses, including the likely effects of drug and lifestyle interventions. Because genetic risk estimates can be calculated at the time of birth, they may provide an early marker for identifying individuals at high risk of developing type 2 diabetes.

Despite this promise, the clinical use of PRSs has been limited. Current scores explain only part of the genetic variance and improve risk prediction only marginally compared with standard clinical risk factors [14,15]. Previous studies have shown that combining multiple PRS may improve predictive accuracy for type 2 diabetes and other diseases compared to using a single PRS alone [16,17]. Importantly, type 2 diabetes is fundamentally a multifactorial disease, and modifiable lifestyle factors (e.g., physical activity and obesity) are key drivers of disease development and progression [18,19]. Genetic variants related to physical activity behavior—such as sedentary behavior (SB) [20]—and traits reflecting physical performance, including grip strength [21,22], cardiorespiratory fitness (CRF) [23], and body mass index (BMI) [24], have been associated with type 2 diabetes and other cardiometabolic diseases [25,26]. These findings suggest that physical activity-related traits are closely intertwined with type 2 diabetes and may further improve genetic prediction.

In light of evidence suggesting potential genetic pleiotropy between type 2 diabetes traits and physical activity behavior [25–27], we propose an expanded approach to genetic prediction. Specifically, we examine whether higher genetic liability for type 2 diabetes is associated with an increased risk of type 2 diabetes and its comorbidities at the population level and whether prediction improves when incorporating physical activity-related genetic liabilities into the model. We hypothesize that including physical activity-related PRSs in addition to the type 2 diabetes PRS will improve the accuracy of predicting type 2 diabetes and its comorbidities.

## METHODS

### Study cohorts

This retrospective cohort study was conducted using FinnGen as the main cohort. It is a public–private partnership research project launched in 2017 that links genotype and digital health record data from Finnish health registries using unique national personal identification numbers (https://www.finngen.fi/en) [28]. We used the FinnGen data release 12 (R12), which includes 520,210 biobank participants (10% of the Finnish population). A complete case approach was applied, requiring participants to have all necessary information for each endpoint analysis. This resulted in a smaller sample size (n = 279,373) compared to the R12.

### Genotyping, quality control, and imputation

The FinnGen individuals were genotyped using Illumina and Affymetrix chip arrays (Illumina Inc., San Diego, and Thermo Fisher Scientific, Santa Clara, CA, USA). Detailed information on genotyping, quality control, and imputation is available in the FinnGen documentation (https://finngen.gitbook.io/documentation/) [28].

### Polygenic risk scoring

Based on a literature search, we selected the publicly available genome-wide association study (GWAS) summary statistics previously published by Mahajan et al. [29] as *the base data* for type 2 diabetes PRS calculation. We selected the most comprehensive publicly available dataset with individuals of European ancestry, as it has been suggested that type 2 diabetes differs in genetic factors and pathophysiology based on ethnicity [30]. The base data included 898,130 genotyped individuals (74,124 type 2 diabetes cases and 824,006 controls) from 32 European ancestry cohorts.

Analysis included several novel, physical activity-related PRSs: physical activity (PA), sedentary behavior (SB), muscular strength (grip), cardiorespiratory fitness (CRF), and body mass index (BMI). All PRSs were based on publicly available GWAS summary statistics from Pan-UK Biobank [32]. With the exception of PA, these PRSs have been previously validated [21,23,33,34]. The PA PRS was validated in this study, with details provided in the Supplemental Material.

Individual genetic scores were calculated using SBayesR, a Bayesian multiple regression approach based on genome-wide summary statistics [35]. SbayesR re-weights summary statistics to account for the linkage disequilibrium between each variant while restricting analyses to HapMap3 SNPs for computational efficiency. All PRSs were genome-wide scores, limited to over 1 million SNPs for computational purposes. The pipeline and code are available elsewhere [21].

### Outcomes

The selection of type 2 diabetes comorbidities was based on the conditions causing the highest healthcare costs in Finland [36–38] and were defined using FinnGen endpoint definitions. Selected outcomes and their ICD-10 codes were type 2 diabetes (E11), hard cardiovascular diseases (hereafter severe cardiovascular diseases [CVD]; I20, I21, I22, I23, I24, I25, I61, I63, I64), diabetic nephropathy (N08.3), diabetic neuropathy (E11.4, G63.2), and diabetic retinopathy (H36.00, H36.02, H36.03). The selected endpoints and their FinnGen Labels are provided in the Supplemental Material. Register data were derived from national hospital discharge records available since 1968. Additional descriptive information, such as disease prevalence and age distribution, can be queried using FinnGen labels via the Risteys interface (https://risteys.finngen.fi/).

### Covariates

Covariates were selected based on prior literature, but their availability in FinnGen was limited. Variables with the most comprehensive data were prioritized. Individual analyses included the first 10 principal components of ancestry, sex, BMI, and smoking status (self-reported yes or no) [39], and genotyping batch, which accounts for the potential stratification that may occur from the genotyping array and sample. Phenotype data were obtained from the Finnish biobanks (https://www.finngen.fi/en/data_protection/data-protection-statement).

### Statistical analysis

Cox proportional hazards models were used to assess associations between PRSs and selected endpoints in the FinnGen cohort. Proportional hazard assumptions were evaluated using Schoenfeld residuals. Potential sex interactions for each PRS were investigated for the type 2 diabetes endpoint, and significant interactions were observed for sex × type 2 diabetes PRS (*p* = 6.9e-04), sex × SB PRS (*p* = 3.0e-04), and sex × BMI PRS (*p* = 2.6e-04). However, the hazard ratios were statistically significant and directionally consistent in both men and women (see Supplemental Material), so analyses were not disaggregated by sex.

Follow-up began at birth for type 2 diabetes outcomes, as PRSs are stable from conception. For comorbidity outcomes, follow-up began at type 2 diabetes diagnosis, ensuring that individuals were only at risk for comorbidities after disease onset and avoiding disease-free survival time before diagnosis, during which the outcome could not have occurred [40]. Follow-up ended at the first occurrence of the endpoint of interest or the end of follow-up, whichever occurred first.

Each PRS was first analyzed individually using Cox regression models. Subsequently, each of the five additional PRSs was added separately to a model to assess incremental predictive value. Finally, all six PRSs were included simultaneously in a multivariable model to evaluate whether combining multiple PRSs improves the prediction beyond type 2 diabetes PRS alone. With > 200 cases per endpoint, sufficient statistical power was expected, using the classical definition of 10 cases per predictor variable.

The concordance index (C-index) was used to evaluate the discriminative ability and performance of the predictive models. The C-index is a measure of a model’s ability to distinguish between individuals who will develop the disease and those who will not [41]. A C-index value of 0.5 indicates no better than random prediction, while a value of 1.0 indicates perfect discrimination.

Standardized PRSs were used in all models; hence, the hazard ratio indicates the risk of incidence per one standard deviation (SD) unit change in the PRS. The significance threshold was set to *p*□< □0.05, with no adjustment for multiple testing. All analyses were performed using R (version 4.4.1, R Foundation for Statistical Computing, Vienna, Austria). We used the R *survival* package [42] to estimate hazard ratios (HRs) and 95% confidence intervals (CIs) between PRSs and incident type 2 diabetes and its comorbidities in the FinnGen cohort.

## RESULTS

Table 1 shows the main characteristics of the FinnGen participants. A slight majority (52.2%) were women, and the mean age at the end of follow-up was 62.0 years (range: 3.5–107.5). Participants’ BMI was 27.4 ± 5.4, and most (73.9%) were non-smokers.

**Table 1.** Descriptives of individuals with selected endpoints in the main FinnGen cohort retinopathy.

| Characteristics | All | n | Women | n | Men | n |
| --- | --- | --- | --- | --- | --- | --- |
| age (y) | 62.0 (17.2) | 279,373 | 59.1 (17.6) | 145,794 | 65.2 (16.2) | 133,579 |
| BMI (kg/m<sup>2</sup>) | 27.4 (5.4) | 279,373 | 27.3 (5.9) | 145,794 | 27.4 (4.7) | 133,579 |
| Smoker n (%) | 73.011 (26.1) | 279,373 | 26.612 (18.3) | 145,794 | 46.399 (34.7) | 133,579 |
| **Comorbidities** |  |  |  |  |  |  |
| T2D | 50.248 (18.0) | 279,373 | 20.890 (14.3) | 145,794 | 29.358 (22.0) | 133,579 |
| CVD severe | 10.073 (23.9) | 42,225 | 15.018 (10.3) | 145,794 | 35.812 (26.8) | 133,579 |
| Diabetic neuropathy | 1.669 (3.4) | 49,652 | 589 (2.8) | 20,798 | 1.410 (4.8) | 29,354 |
| Diabetic nephropathy | 2.345 (4.7) | 49,560 | 861 (4.1) | 20,813 | 1945 (6.6) | 29,377 |
| Diabetic retinopathy | 2.115 (4.6) | 45,558 | 984 (5.1) | 19,182 | 1.536 (5.7) | 26,945 |

Notes: Values are means and standard deviations (SD) for continuous variables, and n and percentages (%) are given for others; BMI, body mass index; T2D, type 2 diabetes; CVD, cardiovascular disease; age at the end of follow-up.

### Associations between PRSs and incident type 2 diabetes

One SD increase in type 2 diabetes PRS was associated with an 8% higher risk of incident type 2 diabetes (HR 1.08, [95% CI 1.08–1.08]; Table 2). All physical activity-related PRSs were associated with type 2 diabetes risk. Lower risk was observed for PA 7% (HR 0.93, [95% CI 0.92–0.93]), grip 4% (HR 0.96, [95% CI 0.95–0.96]), and CRF 6% (HR 0.94, [95% CI 0.94–0.95]). Higher risk was observed for SB 14% (HR 1.14, [95% CI 1.13–1.15]) and BMI 35% (HR 1.35, [95% CI 1.34–1.36]). However, these associations were slightly attenuated when the models were adjusted for measured BMI and smoking status (Table 2).

**Table 2.** Associations between PRSs and T2D incidence.

| T2D | Model 1<sup>a</sup> |  | Model 2<sup>b</sup> |  |
| --- | --- | --- | --- | --- |
|  | HR (95% CI) | p Value | HR (95% CI) | p Value |
| **PRS** |  |  |  |  |
| T2D | 1.08 (1.08–1.08) | **0.0e+00** | 1.07 (1.07–1.08) | **0.0e+00** |
| PA | 0.93 (0.92–0.93) | **1.2e-67** | 0.95 (0.94–0.96) | **4.4e-30** |
| SB | 1.14 (1.13–1.15) | **4.4e-197** | 1.09 (1.08–1.10) | **4.7e-78** |
| Grip | 0.96 (0.95–0.96) | **5.7e-24** | 0.96 (0.95–0.96) | **2.6e-24** |
| CRF | 0.94 (0.94–0.95) | 1.3e-36 | 0.96 (0.96–0.97) | 7.2e-16 |
| BMI | 1.35 (1.34–1.36) | **0.0e+00** | 1.13 (1.12–1.14) | **2.5e-144** |
| T2D + PA | 1.08 (1.08–1.08) | **0.0e+00** | 1.07 (1.07–1.08) | **0.0e+00** |
| T2D + SB | 1.08 (1.07–1.08) | **0.0e+00** | 1.07 (1.07–1.08) | **0.0e+00** |
| T2D + Grip | 1.08 (1.08–1.08) | **0.0e+00** | 1.07 (1.07–1.08) | **0.0e+00** |
| T2D + CRF | 1.08 (1.08–1.08) | **0.0e+00** | 1.07 (1.07–1.08) | **0.0e+00** |
| T2D + BMI | 1.07 (1.07–1.08) | **0.0e+00** | 1.07 (1.07–1.08) | **0.0e+00** |
| All PRSs | 1.07 (1.07–1.08) | **1.5e-320** | 1.07 (1.07–1.08) | **0.0e+00** |

a. Model 1 adjusted for sex, age, 10 genetic principal components, and genotyping batch.
b. Model 2 adjusted for sex, age, 10 genetic principal components, genotyping batch, BMI, and smoking status. PRSs: T2D, Type 2 diabetes; PA, physical activity; Grip, handgrip; CRF cardiorespiratory fitness; BMI, body mass index.

### Associations between PRSs and incident type 2 diabetes comorbidities

#### Diabetic nephropathy

One SD increase in type 2 diabetes PRS was associated with a 4% higher risk of diabetic nephropathy (HR 1.04, [95% CI 1.02–1.06]; Table 3). PA and CRF PRSs were associated with a lower risk of nephropathy, while SB and BMI PRSs were linked to a higher risk.

**Table 3.**
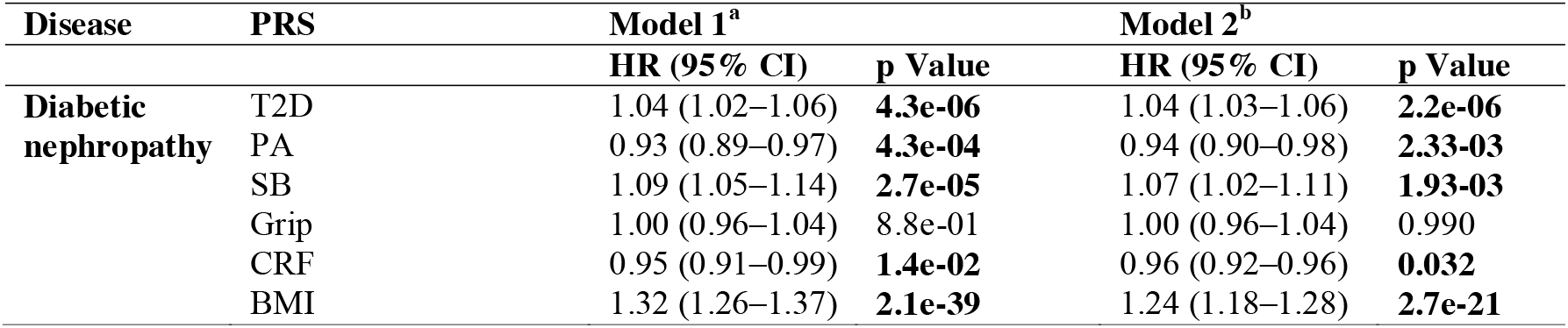
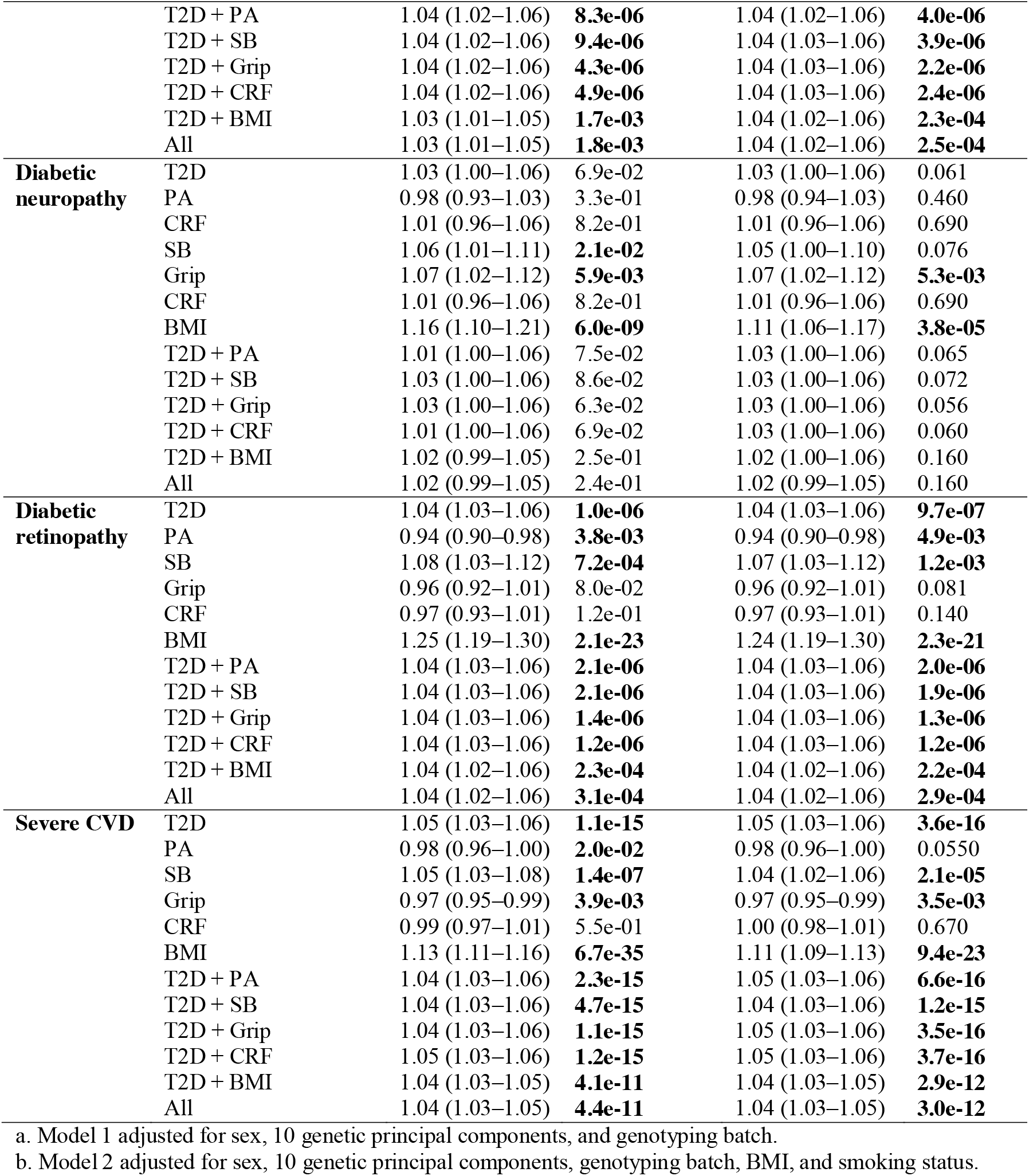
Associations between PRSs and T2D comorbidities.

| Disease | PRS | Model 1<sup>a</sup> |  | Model 2<sup>b</sup> |  |
| --- | --- | --- | --- | --- | --- |
|  |  | HR (95% CI) | p Value | HR (95% CI) | p Value |
| Diabetic nephropathy | T2D | 1.04 (1.02–1.06) | **4.3e-06** | 1.04 (1.03–1.06) | **2.2e-06** |
|  | PA | 0.93 (0.89–0.97) | **4.3e-04** | 0.94 (0.90–0.98) | **2.33-03** |
|  | SB | 1.09 (1.05–1.14) | **2.7e-05** | 1.07 (1.02–1.11) | **1.93-03** |
|  | Grip | 1.00 (0.96–1.04) | 8.8e-01 | 1.00 (0.96–1.04) | 0.990 |
|  | CRF | 0.95 (0.91–0.99) | **1.4e-02** | 0.96 (0.92–0.96) | **0.032** |
|  | BMI | 1.32 (1.26–1.37) | **2.1e-39** | 1.24 (1.18–1.28) | **2.7e-21** |
|  | T2D + PA | 1.04 (1.02–1.06) | **8.3e-06** | 1.04 (1.02–1.06) | **4.0e-06** |
|  | T2D + SB | 1.04 (1.02–1.06) | **9.4e-06** | 1.04 (1.03–1.06) | **3.9e-06** |
|  | T2D + Grip | 1.04 (1.02–1.06) | **4.3e-06** | 1.04 (1.03–1.06) | **2.2e-06** |
|  | T2D + CRF | 1.04 (1.02–1.06) | **4.9e-06** | 1.04 (1.03–1.06) | **2.4e-06** |
|  | T2D + BMI | 1.03 (1.01–1.05) | **1.7e-03** | 1.04 (1.02–1.06) | **2.3e-04** |
|  | All | 1.03 (1.01–1.05) | **1.8e-03** | 1.04 (1.02–1.06) | **2.5e-04** |
| **Diabetic neuropathy** | T2D | 1.03 (1.00–1.06) | 6.9e-02 | 1.03 (1.00–1.06) | 0.061 |
|  | PA | 0.98 (0.93–1.03) | 3.3e-01 | 0.98 (0.94–1.03) | 0.460 |
|  | CRF | 1.01 (0.96–1.06) | 8.2e-01 | 1.01 (0.96–1.06) | 0.690 |
|  | SB | 1.06 (1.01–1.11) | **2.1e-02** | 1.05 (1.00–1.10) | 0.076 |
|  | Grip | 1.07 (1.02–1.12) | **5.9e-03** | 1.07 (1.02–1.12) | **5.3e-03** |
|  | CRF | 1.01 (0.96–1.06) | 8.2e-01 | 1.01 (0.96–1.06) | 0.690 |
|  | BMI | 1.16 (1.10–1.21) | **6.0e-09** | 1.11 (1.06–1.17) | **3.8e-05** |
|  | T2D + PA | 1.01 (1.00–1.06) | 7.5e-02 | 1.03 (1.00–1.06) | 0.065 |
|  | T2D + SB | 1.03 (1.00–1.06) | 8.6e-02 | 1.03 (1.00–1.06) | 0.072 |
|  | T2D + Grip | 1.03 (1.00–1.06) | 6.3e-02 | 1.03 (1.00–1.06) | 0.056 |
|  | T2D + CRF | 1.01 (1.00–1.06) | 6.9e-02 | 1.03 (1.00–1.06) | 0.060 |
|  | T2D + BMI | 1.02 (0.99–1.05) | 2.5e-01 | 1.02 (1.00–1.06) | 0.160 |
|  | All | 1.02 (0.99–1.05) | 2.4e-01 | 1.02 (0.99–1.05) | 0.160 |
| **Diabetic retinopathy** | T2D | 1.04 (1.03–1.06) | **1.0e-06** | 1.04 (1.03–1.06) | **9.7e-07** |
|  | PA | 0.94 (0.90–0.98) | **3.8e-03** | 0.94 (0.90–0.98) | **4.9e-03** |
|  | SB | 1.08 (1.03–1.12) | **7.2e-04** | 1.07 (1.03–1.12) | **1.2e-03** |
|  | Grip | 0.96 (0.92–1.01) | 8.0e-02 | 0.96 (0.92–1.01) | 0.081 |
|  | CRF | 0.97 (0.93–1.01) | 1.2e-01 | 0.97 (0.93–1.01) | 0.140 |
|  | BMI | 1.25 (1.19–1.30) | **2.1e-23** | 1.24 (1.19–1.30) | **2.3e-21** |
|  | T2D + PA | 1.04 (1.03–1.06) | **2.1e-06** | 1.04 (1.03–1.06) | **2.0e-06** |
|  | T2D + SB | 1.04 (1.03–1.06) | **2.1e-06** | 1.04 (1.03–1.06) | **1.9e-06** |
|  | T2D + Grip | 1.04 (1.03–1.06) | **1.4e-06** | 1.04 (1.03–1.06) | **1.3e-06** |
|  | T2D + CRF | 1.04 (1.03–1.06) | **1.2e-06** | 1.04 (1.03–1.06) | **1.2e-06** |
|  | T2D + BMI | 1.04 (1.02–1.06) | **2.3e-04** | 1.04 (1.02–1.06) | **2.2e-04** |
|  | All | 1.04 (1.02–1.06) | **3.1e-04** | 1.04 (1.02–1.06) | **2.9e-04** |
| **Severe CVD** | T2D | 1.05 (1.03–1.06) | **1.1e-15** | 1.05 (1.03–1.06) | **3.6e-16** |
|  | PA | 0.98 (0.96–1.00) | **2.0e-02** | 0.98 (0.96–1.00) | 0.0550 |
|  | SB | 1.05 (1.03–1.08) | **1.4e-07** | 1.04 (1.02–1.06) | **2.1e-05** |
|  | Grip | 0.97 (0.95–0.99) | **3.9e-03** | 0.97 (0.95–0.99) | **3.5e-03** |
|  | CRF | 0.99 (0.97–1.01) | 5.5e-01 | 1.00 (0.98–1.01) | 0.670 |
|  | BMI | 1.13 (1.11–1.16) | **6.7e-35** | 1.11 (1.09–1.13) | **9.4e-23** |
|  | T2D + PA | 1.04 (1.03–1.06) | **2.3e-15** | 1.05 (1.03–1.06) | **6.6e-16** |
|  | T2D + SB | 1.04 (1.03–1.06) | **4.7e-15** | 1.04 (1.03–1.06) | **1.2e-15** |
|  | T2D + Grip | 1.04 (1.03–1.06) | **1.1e-15** | 1.05 (1.03–1.06) | **3.5e-16** |
|  | T2D + CRF | 1.05 (1.03–1.06) | **1.2e-15** | 1.05 (1.03–1.06) | **3.7e-16** |
|  | T2D + BMI | 1.04 (1.03–1.05) | **4.1e-11** | 1.04 (1.03–1.05) | **2.9e-12** |
|  | All | 1.04 (1.03–1.05) | **4.4e-11** | 1.04 (1.03–1.05) | **3.0e-12** |

a. Model 1 adjusted for sex, 10 genetic principal components, and genotyping batch.
b. Model 2 adjusted for sex, 10 genetic principal components, genotyping batch, BMI, and smoking status.

#### Diabetic retinopathy

Type 2 diabetes PRS was associated with a 4% higher risk of diabetic retinopathy (HR 1.04, [95% CI 1.03 to 1.06]; Table 3). PA PRS predicted a lower risk of retinopathy, while SB and BMI PRSs predicted a higher risk.

#### Diabetic neuropathy

The type 2 diabetes PRS was not significantly associated with diabetic neuropathy. SB and BMI PRSs predicted higher risk, while grip PRS was also linked to an increased risk of neuropathy.

#### Severe CVD

Type 2 diabetes PRS was associated with a 5% higher risk of severe CVD (HR 1.05 [95% CI 1.03 to 1.06]). PA, grip, and CRF PRSs were linked to a modestly reduced risk of CVD (1–3%), whereas SB and BMI PRSs were associated with a higher risk of CVD. After adjusting the model for measured BMI and smoking status, PA PRS was no longer statistically significant.

### Prediction of type 2 diabetes and comorbidities with PRSs

Model performance was also evaluated using the concordance index. In the base model, which incorporated only the type 2 diabetes PRS for risk prediction, the C-index was 0.644. Adding other PRSs had little effect, with a maximum increase of 0.028, and the HR for type 2 diabetes PRS remained largely unchanged. In contrast, adding measured BMI and smoking status increased the C-index for type 2 diabetes by 0.1, but not for comorbidities (Table 4).

**Table 4.**
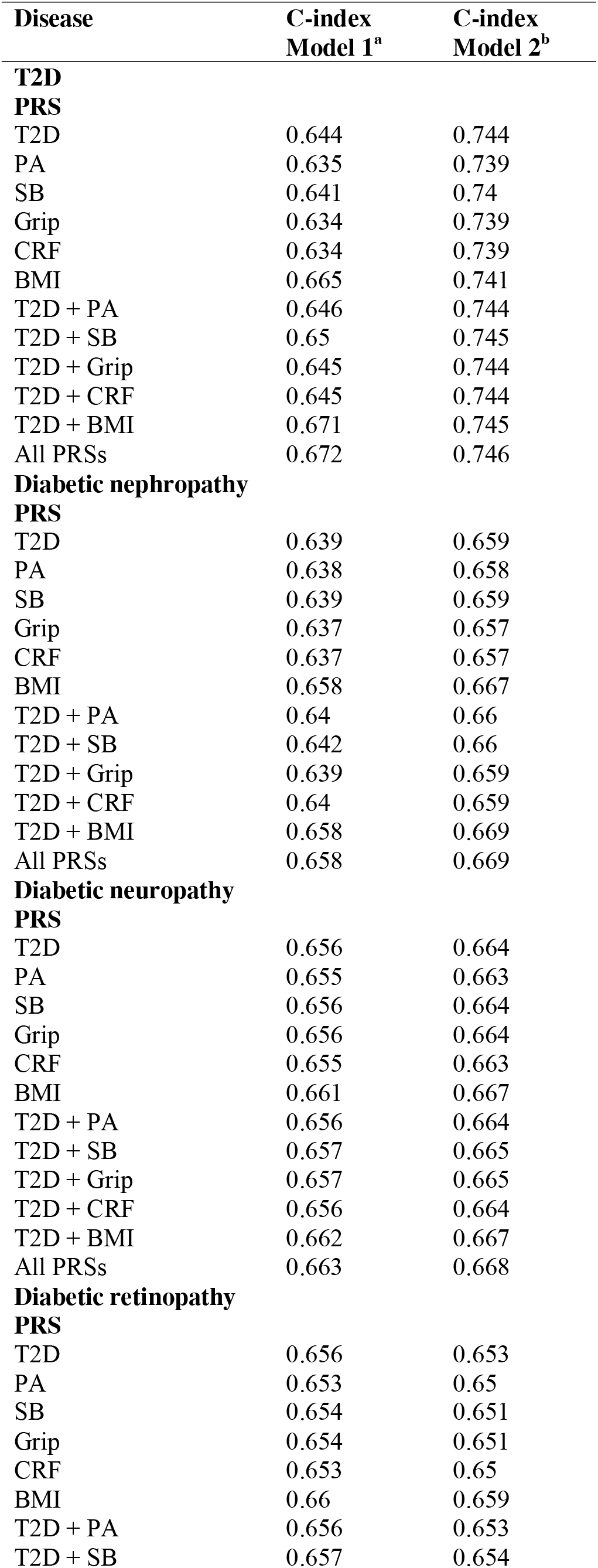
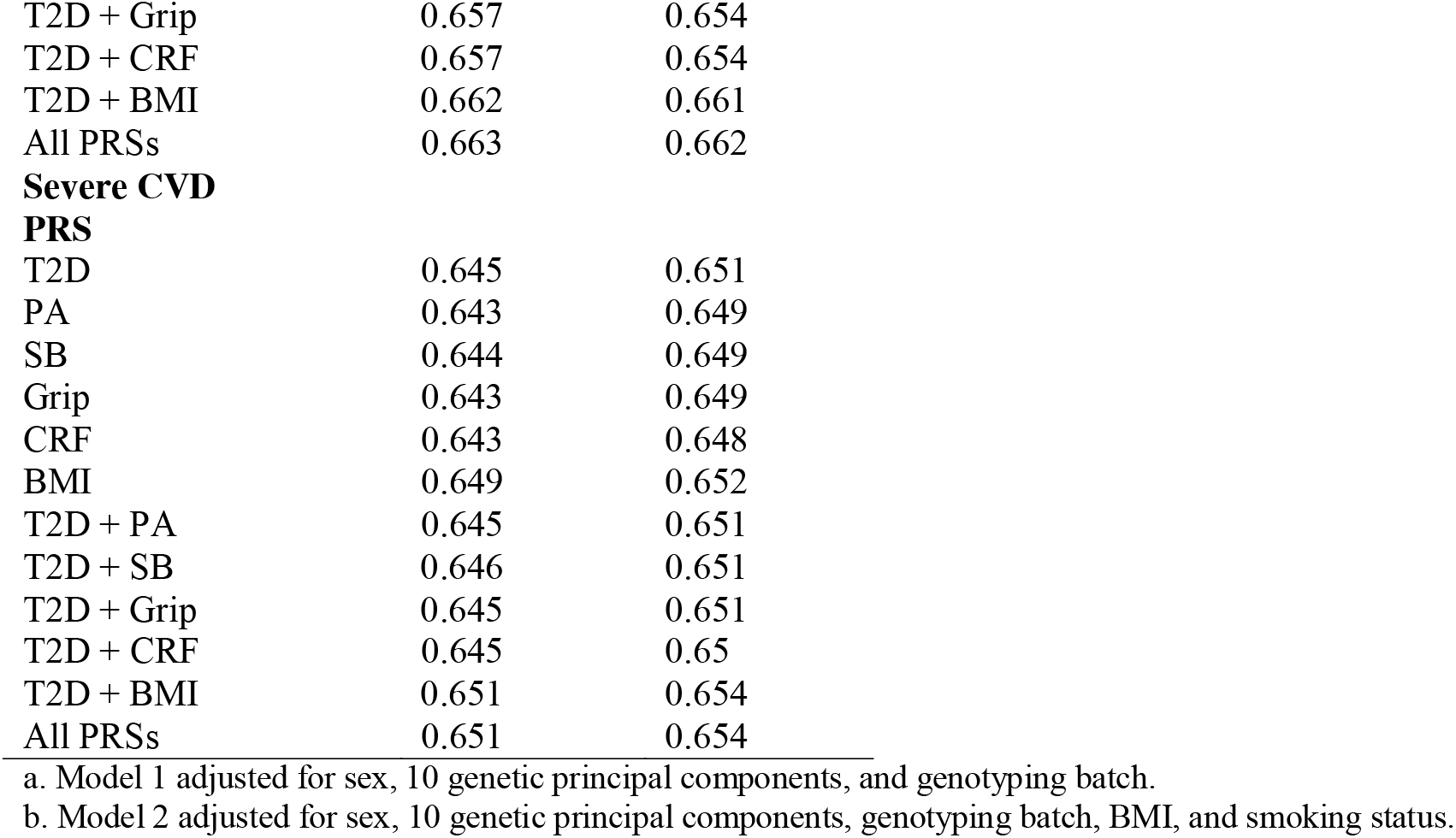
PRSs as a Predictor of T2D in FinnGen.

## DISCUSSION

The aim of this study was to assess how genetic factors, including those related to physical activity, contribute to the risk of developing type 2 diabetes and its comorbidities in the Finnish population. We found that higher genetic liability to type 2 diabetes was associated with a greater risk of developing the disease, consistent with previous studies [12,43]. Among individuals with type 2 diabetes, type 2 diabetes PRS was associated with a higher risk of nephropathy, retinopathy, and severe CVDs, but not with neuropathy. Several physical activity-related PRSs were independently associated with type 2 diabetes and its comorbidities, but their inclusion in the model did not significantly improve prediction, suggesting potential genetic pleiotropy.

To the best of our knowledge, this is the first study to examine whether physical activity-related polygenic scores can improve the prediction of type 2 diabetes. While previous research has examined interactions between lifestyle and genetic risk, no studies have directly combined physical activity-related PRSs with type 2 diabetes PRS in a single predictive framework. Although adding physical activity-related PRSs did not improve predictive accuracy in our study, this approach provides a novel perspective. It may support the hypothesis that shared genetic factors influence both type 2 diabetes susceptibility and physical activity behavior. This aligns with earlier findings suggesting that physical activity is heritable and genetically correlated with several cardiometabolic traits, including type 2 diabetes [25,44]. Grip strength—a proxy for muscle strength—has been associated with improved metabolic health and reduced type 2 diabetes risk, likely through mechanisms involving greater muscle mass and enhanced insulin sensitivity [21,22]. Likewise, genetic predisposition to sedentary behavior is correlated with increased type 2 diabetes risk, supporting the role of prolonged inactivity in disease development [45]. Similarly, BMI is a well-established risk factor, with higher BMI consistently associated with an increased risk of type 2 diabetes [24].

The limited improvement in prediction, as indicated by the C-index, could be due to overlapping genetic signals between PRSs, their low individual explanatory power, and the complex polygenic architecture of type 2 diabetes and related traits [29,46]. Nevertheless, methodological advances may improve the integration of polygenic scores in the future. Notably, predictive performance improved more clearly when measured lifestyle-related phenotypic variables, such as smoking and BMI, were added to the model (C-index increased from 0.644 to 0.744), compared to the addition of physical activity-related PRSs. This suggests that direct measures of lifestyle factors currently add more predictive value than their polygenic proxies, although methodological development may narrow this gap.

Although the observed effect sizes were modest, our findings support the potential role of PRSs in preventive strategies for type 2 diabetes. Even modest gains in risk stratification may have clinical significance, especially in terms of early intervention and targeted prevention.

Currently, the clinical use of PRSs is largely confined to research and pilot programs, but their potential for disease prevention is increasingly recognized. Traditional risk assessment tools remain essential for population-level screening, yet our findings suggest that PRSs could provide an additional, genetically informed layer of risk evaluation. Recent studies also indicate that PRS-based screening may be cost-effective when focused on high-risk individuals and combined with affordable genotyping technologies [47]. Importantly, observational data show that individuals with high polygenic risk still benefit significantly from healthy lifestyle choices, underscoring the value of early risk identification [48]. Thus, while PRS-based prevention is not yet part of standard care, it may hold promise as a future tool for more personalized and proactive disease prevention strategies.

Considering the inclusion of multiple validated PRSs in this study, it is noteworthy that while all physical activity-related PRSs predicted T2D risk to some extent, their predictive performance varied. These differences may partly reflect disparities in the underlying GWAS data used to construct the scores. For instance, the PRS for cardiorespiratory fitness was derived from a considerably smaller GWAS sample compared to those for sedentary behavior or BMI, potentially limiting its predictive power due to reduced statistical power and variant discovery. Sedentary behavior and BMI are well-established lifestyle-related risk factors for type 2 diabetes, which may explain their stronger predictive performance. These traits are also likely to have more robust and well-powered GWASs, resulting in more informative PRSs. We found that the BMI PRS outperformed the type 2 diabetes PRS in predicting type 2 diabetes and its comorbidities. This suggests that PRSs for intermediate traits, such as BMI, may outperform disease-specific PRSs in predicting future disease onset. Our findings are consistent with UK Biobank analyses showing that the BMI PRS predicted type 2 diabetes incidence more accurately than the type 2 diabetes PRS, particularly in women [49]. Recent research further supports the utility of PRSs in identifying individuals at elevated genetic risk before the onset of clinical symptoms [50]. Based on the current evidence, PRSs reflecting modifiable risk pathways may capture important biological mechanisms underlying disease development and offer greater utility for screening.

### Strengths and limitations

This study has several strengths. We applied high-performing, genome-wide methods for genetic scoring. Specifically, we used the SBayesR method to derive type 2 diabetes PRS, which has been shown to outperform other commonly utilized PRS methods. A key strength is the use of GWAS data from Mahajan et al. (2018) [29], which provide the best predictive performance for type 2 diabetes PRSs when combined with advanced Bayesian methods such as SBayesR [31]. Another strength is the inclusion of multiple validated PRSs, including the most up-to-date and best-performing scores for physical activity-related traits, thereby enhancing the robustness of genetic risk assessment. Furthermore, we leveraged the large-scale FinnGen cohort, which covers approximately 10% of the Finnish adult population, and linked genetic data to comprehensive national health registers. This enabled detailed and reliable phenotyping with near-complete follow-up, which strengthens the reliability of our findings. To the best of our knowledge, FinnGen has not been widely used in previous studies of type 2 diabetes PRSs, making this study a valuable addition to the literature by validating findings in an independent, population-based dataset and broadening the generalizability of PRS research beyond commonly used cohorts.

However, the generalizability of our findings is limited to European populations, as both the underlying GWAS data and the FinnGen cohort are composed of individuals of European descent. Our analyses were also constrained by the lack of detailed covariate data, particularly on lifestyle factors such as diet, physical activity, and socioeconomic status. Nonetheless, the primary aim of this study was to evaluate the independent predictive utility of type 2 diabetes PRSs in early-stage screening, rather than to test their performance when adjusted for lifestyle-related variables. Additionally, FinnGen may overrepresent individuals with a higher disease burden, as participants are primarily recruited from those with healthcare contact, which could influence the generalizability of lifetime risk estimates.

## Conclusions

PRSs for type 2 diabetes and physical activity-related phenotypes independently predict the incidence of type 2 diabetes and related comorbidities. However, adding physical activity-related scores to the model did not meaningfully improve prediction. These findings support the hypothesis that genetic pleiotropy may partly explain associations between type 2 diabetes and physical activity behavior.

## Data Availability

Data are available upon reasonable request. Researchers can apply to use the FinnGen resource and access the data used. The Finnish biobank data can be accessed through the Fingenious services (https://site.fingenious.fi/en/) managed by FINBB. The FTC subsample data are taken from the Biobank of the National Institute for Health and Welfare. Data are available for qualified researchers through a standardized application procedure.

## Acknowledgements

We thank the participants and all those who contributed to the data collection in the FTC and FinnGen cohorts, without whom this study would not have been possible. A detailed list of investigators involved in FinnGen is provided in the Supplemental Material (pages 2–17). We used AI-assisted tools (DeepL and ChatGPT) for language editing, proofreading, and improving the clarity and structure of the manuscript. All content was reviewed and verified by the authors.

## Abbreviations

CRF: Cardiorespiratory fitness
GWAS: Genome-wide association study
PA: Physical activity
PRS: Polygenic risk score
SB: Sedentary behavior

## Disclosures

### Data availability

Data are available upon reasonable request. Researchers can apply to use the FinnGen resource and access the data used. The Finnish biobank data can be accessed through the Fingenious services (https://site.fingenious.fi/en/) managed by FINBB (Finnish Biobank Cooperative – FINBB | Finnish Biobank Cooperative – FINBB). The FTC subsample data are taken from the Biobank of the National Institute for Health and Welfare. Data are available for qualified researchers through a standardized application procedure (see details Data and services - THL).

### Contributorship

EV, LJ, KW, and ES conceptualized the research question, study design, and statistical analysis. EV and LJ preprocessed the polygenic score data and implemented the scores to the FinnGen study cohort. EV performed the statistical modelling under the supervision of LJ and ES. EV drafted the first version of the manuscript, and all authors contributed significantly to the writing, critical interpretation of the findings, and revision of the manuscript. The corresponding author attests that all listed authors meet authorship criteria and that no others meeting the criteria have been omitted.

